# Services in Minoritized Autistic Adolescents and Adults Varying in Language Skills

**DOI:** 10.1101/2024.05.26.24307944

**Authors:** Teresa Girolamo, Alicia Escobedo, Samantha Ghali, Kyle Greene-Pendelton, Iván Campos, Poornima Ram-Kiran

## Abstract

**Background:** Racially and ethnically minoritized (minoritized) autistic individuals face intersectional disparities in services access in the transition to adulthood. Our understanding of disparities is limited by systematic exclusion from research and inadequate approaches to characterizing services. To address these gaps and effect advocacy, this study: 1) examined services received, unmet service needs, and barriers in minoritized autistic adolescents and adults, and 2) determined if language, NVIQ, and autism traits predict services when deployed as binary or continuous variables.

**Method:** Academic and community partners tailored CBPR to a local context. Participants (*N* = 73, ages 13-30) completed a behavioral assessment protocol. Participants and caregivers provided information on services received, unmet service needs, and barriers to services. Data were analyzed using descriptives and regression.

**Results:** Participants received multiple services yet had multiple unmet service needs and barriers. Effects of services differed by approach. Language impairment, but not language scores, predicted receiving more services. High levels of autism traits and autism trait scores predicted more unmet service needs.

**Implications:** While the number of services and unmet service needs were similar to prior work, differences in individual service variables and effects support attention to heterogeneity. Findings support intersectional approaches to CBPR and autism research.

**Learning outcomes:** After reading the article, the learner will be able to: 1) summarize knowledge gaps about access to services; 2) explain the relevance of tailoring CBPR to a local context; and 3) describe implications of findings for clinicians and autistic individuals.

---

In the United States (U.S.), autism has a diagnosed prevalence that is higher in Black, Hispanic, and Asian and Pacific Islander youth than white youth (1 in 30-34 versus 1 in 42) (Maenner et al., 2023). While social communication is central to an autism diagnosis (American Psychiatric Association [APA], 2013; World Health Organization [WHO], 2022), over 50% of autistic individuals have structural language impairment (LI; Boucher, 2012), or challenges in phonology, morphology, and syntax (Schaeffer et al., 2023). Yet autistic individuals who are racially and ethnically minoritized (hereafter, minoritized) and those with LI face intersectional disparities in adult outcomes (Johnson et al., 2010; Schott et al., 2022) and systematic exclusion from research (Girolamo et al., 2023a). Systematic reviews on evidence-based autism intervention (Steinbrenner et al., 2022) and assessment of structural language in autism (Girolamo et al., 2023a) revealed 63.6% and 77% of participants (ages birth to 22 and 3 to 21), respectively, were white; 9.4% and 7.9% Hispanic or Latine; 7.7% and 6.2% Black; and 6.4% and 0.7% Asian or Pacific Islander (Steinbrenner et al., 2022). Currently, 58.4% of the U.S. population is white, 19.5% Hispanic or Latine, 13.7% Black, and 6.4% Asian, (United States Census Bureau, 2024). Thus, even with growing attention to services in the transition to adulthood (“Individuals with Disabilities Education Improvement Act [IDEIA] of 2004,” 2018; Roux et al., 2015), a nonrepresentative evidence base limits the ability to reduce disparities (National Institutes of Health, 2021). With over 1.2 million autistic youth in entering adulthood in the next decade (Interagency Autism Coordinating Committee, 2023), a broad understanding of services in autistic individuals is crucial for health equity.

Though under-utilized in autism research (Chen et al., 2024), community-based participatory research (CBPR) is a proven way to reduce disparities (Wallerstein & Duran, 2017) and aligns with evidence-based practice: integrating expert opinion, individual preference, and best-available evidence (Sackett et al., 1996). In using full partnership with shared power to generate knowledge for change (Wallerstein & Duran, 2006, 2010), CBPR is at the middle of a community-engaged research model continuum that ranges from lesser (e.g., communities inform, consult, participate in, or initiate research) to greater community engagement and power (e.g., communities lead and own research; Key et al., 2019). CBPR is guided by principles: a) recognize the community as a unit of identity; b) address complex contexts, such as cultural factors and historical mistrust; c) use a social-ecological approach, which attributes experiences of interactions between individuals and multi-level environmental influences (Bronfenbrenner, 1977), from family (e.g., household income) and interpersonal (e.g., social networks) to institutional (e.g., participation in institutions, such as schools), community (e.g., availability of services), and societal (e.g., policy for service eligibility; Anderson et al., 2018b; National Institute on Minority Health and Health Disparities [NIMHD], 2017); d) acknowledge and build upon community strengths (Clinical and Translational Science Awards Consortium [CTSA], 2011); e) foster equitable partnership in all research phases, given individual, relational, and structural power dynamics; f) balance knowledge generation with action to benefit all partners; g) disseminate findings with partners to the community; h) commit to sustained partnership to effect change; and i) engage in a reflective, cyclical process (Israel et al., 2003, 2010; Wallerstein et al., 2008). In all, CBPR is a holistic framework ideal for studying services for advocacy.

However, prior work reveals inadequate attention to intersectional nuance (Anderson et al., 2018b; Chen et al., 2024; Fletcher-Watson et al., 2019; Nicolaidis & Raymaker, 2015), which is critical in considering the experiences of transition-aged autistic individuals (Schendel et al., 2022). Theories from special education (Annamma et al., 2013), legal history (Crenshaw, 1989, 1991), and psychology (Plaut, 2010) frame race and disability as mutually reinforcing social constructs that cause social marginalization. That is, perceptions of others about race and disability versus minoritized autistic individuals themselves lead to racism and ableism (Annamma et al., 2016). For example, U.S. law enacted race- and disability-based segregation in education; later policy to desegregate schools involved changes in societal norms and not in students (Powell, 2012; Turnbull III & Turnbull, 1998). Persisting intersectional disparities in access to services similarly implicate social processes (Annamma, 2018; Annamma et al., 2013; Kasambira Fannin et al., 2024). Minoritized families of autistic youth report reduced access to services due to practitioners lacking knowledge about community norms (Onovbiona et al., 2023; Weitlauf et al., 2024), as well as greater autism-related stigma than white families (Rivera-Figueroa et al., 2022). At the same time, there is no one-to-one ratio between autism, race, and experience. While autistic adults report stigma from disclosure (Thompson-Hodgetts et al., 2020), Black autistic adults report racism that can intersect with ableism (Davis et al., 2024). Understanding heterogeneity in the experiences of minoritized individuals is important.

A broader question is how to apply CBPR to study services in minoritized autistic individuals. Suggested practices to enhance inclusion in autism research are diversity advisory boards (Williams et al., 2023) and community involvement statements (Autism, 2024). In requiring disclosure, each risks perpetuating inequity (Wallerstein et al., 2019). For instance, a journal may pressure a minoritized autistic author of a manuscript to disclose their diagnosis in order to “prove” inclusion (Oswald, 2024), without any guarantee of confidentiality or nondiscrimination. In turn, a white editorial board member who reviews the manuscript may violate journal policies and professional ethics by sharing private information about the author with a therapist network. If the journal fails to act upon learning of these violations, it amounts to diversity dishonesty and violates CBPR principles (Wilton et al., 2020). Given these considerations, this report examines services in minoritized autistic adolescents and adults varying in language skills and shares use of CBPR.

## Services in Minoritized Autistic Adolescents and Adults

The best-available evidence shows variability in services received, unmet service needs, and barriers to services in transition-aged autistic individuals. The National Longitudinal Transition Survey 2 (NLTS2) followed a nationally representative sample of >11,000 adolescents receiving special education services from 2000 to 2010 and assessed receipt of 16 services per special education legislation (“Individuals with Disabilities Education Act [IDEA] of 1997,” 1997) and barriers to services (Levine et al., 2007); see Supplementary Table 1 for full list. In high school, 74.6% of NLTS2 youth (ages 13 to 18) received speech-language therapy, and other service utilization rates ranged from 2.5% for audiology to 54.6% for transportation (Levine et al., 2007). Nearly 40% of 410 NLTS2 youth (ages 19 to 23) received no services post-high school, and 9.1% received speech-language therapy (Shattuck et al., 2011). Subsequent studies referencing NLTS2 services had mixed findings; see Supplementary Tables 1 to 3. For example, 98.2% of 168 autistic individuals (ages 16 to 30, 60% post-secondary) received at least one service, but the number of services received (*M*: 6.13, 0-15; Ishler et al., 2022) and unmet service needs (*M*: 3.18, 0-11) varied (Ishler et al., 2023); see also, Taylor and Henninger (2015). Interpreting this variability is challenging for several reasons.

First, studies examined racial disparities in services using methods that erased heterogeneity, despite broad patterns showing no one-to-one ratio between services and race (Eilenberg et al., 2019; Liu et al., 2023; Schott et al., 2021). Some studies found racial disparities in services received and unmet needs (Shattuck et al., 2011; Taylor & Henninger, 2015), while others did not (Ishler et al., 2022, 2023). These findings are based on group comparisons of white and minoritized groups, with 77% to 93.6% white samples (Ishler et al., 2022, 2023; Laxman et al., 2019; Song et al., 2022; Taylor & Henninger, 2015; Turcotte et al., 2016). In some cases, analysis excluded Asian/Pacific Islander, Latino, Hispanic, or Chicano, and Native American participants (Turcotte et al., 2016), or “adjusted” for race (Song et al., 2022). These practices erase variance (Swilley-Martinez et al., 2023). Studies also used inconsistent definitions for “minoritized.” Reports on the same sample defined “underrepresented minority” as Black, Hispanic, or multiracial (Ishler et al., 2022) and “historically marginalized minority” as also including Native American or Alaska Native (Ishler et al., 2023). Understanding services patterns requires moving beyond white-minoritized comparisons.

A second limitation was characterizing language and cognitive skills with confounding approaches. Shattuck et al. (2011) found no effects of nonverbal status on services received but equated “nonverbal” with “severe LI,” or caregiver report of “has a lot of trouble speaking clearly” or “doesn’t speak at all.” This suggests spoken language entails linguistic knowledge. In reality, language profiles in autism vary by domain and definition (Bal et al., 2016; Butler et al., 2023; Girolamo et al., 2023b; Koegel et al., 2020). “Speaking clearly” could involve motor speech (Shriberg et al., 2019), lexical access (Arunachalam & Luyster, 2016), or morphosyntax (Sterling, 2018). To strengthen characterization of LI, assessment across domains is needed. Studies also examined intellectual disability (ID), finding no effects on services received, unmet needs, and barriers (Ishler et al., 2023; Ishler et al., 2022; Laxman et al., 2019; Taylor & Henninger, 2015) or positive effects on services received (Laxman et al., 2019). However, studies determined ID using measures that include language (Laxman et al., 2019; Sparrow et al., 1993; Sparrow et al., 2005) or require verbal language to complete tasks (Roid & Pomplun, 2003; Taylor & Henninger, 2015). Language and IQ can dissociate in autism (Manenti et al., 2024; Silleresi et al., 2020), even when considering autism traits (Bal et al., 2016). Also, IQ measures requiring verbal language yield scores nearly 1 *SD* lower than NVIQ measures in autistic youth with communication age equivalents below 4.5 years (Grondhuis et al., 2018). While LI and ID are clinically relevant, a dimensional approach to characterizing language skills across domains and NVIQ is needed (Kover & Abbeduto, 2023).

Third, amid a broad knowledge gap on factors in outcomes of older autistic individuals (Schendel et al., 2022), studies overlooked interpersonal social-ecological factors. Some studies focused on high school enrollment, an institutional factor (Anderson et al., 2018b). High school exit was tied to receiving fewer services and greater unmet service needs (Laxman et al., 2019; Song et al., 2022; Turcotte et al., 2016). Elsewhere, high school enrollment predicted receiving more services but not barriers (Ishler et al., 2022, 2023). Amid calls to focus on the social consequences of race and disability in minoritized autistic individuals (Davis et al., 2024), a promising interpersonal factor is sense of community (Peterson et al., 2008). As a proxy for social capital, sense of community is linked to network-based resources (Carpiano & Hystad, 2011; Michalski et al., 2020), as well as access to services in autistic youth (Gulsrud et al., 2021). In turn, barriers to services are linked to community access, which may limit sense of community (Cameron et al., 2022). While sense of community can be reliably and sensitively measured in Black and Hispanic adolescents (Lardier Jr et al., 2018, 2022; Opara et al., 2021) and adults from the general population (Cardenas et al., 2021), it has yet to be investigated in autism research.

## The Current Study

Evidence to date points to the need for nuance in use of CBPR to study services in minoritized autistic individuals in the transition to adulthood. Given theories to frame race and disability (Annamma et al., 2013; Crenshaw, 1989, 1991; Plaut, 2010) and approaches to characterize heterogeneity in autism (Kover & Abbeduto, 2023; Schaeffer et al., 2023), this study aimed to generate knowledge for advocacy. A focus was multi-level influences on services, using direct assessment to characterize language skills across domains and NVIQ. Last, use of CBPR aligns with evidence-based practice (Burns et al., 2011). This study asked:

1. What are patterns in services received, unmet service needs, and barriers to having service need met in minoritized autistic adolescents and adults?
2. To what extent are services received, unmet service needs, and barriers to services, predicted by individual language skills and NVIQ, educational enrollment, and sense of community, and how do effects compare when using a categorical versus continuous approach to language and NVIQ?

## Method

This study received institutional board approval and followed all ethical guidelines.

### Participatory Approach

This study began as discussions with 5 minoritized autistic adults, 3 caregivers, and practitioners (3 speech-language pathologists, 2 psychologists, 2 special education teachers) about disparities in research and practice based on their experiences. A research team member had met a self-advocate at an event for minoritized professionals, self-advocates, and families, connected over shared interests in intersectional approaches to evidence-based practice, and the self-advocate invited others to their discussions. The aim was to address community priorities, such as community-based assessment to provide access to services to minoritized autistic youth and adults who were being denied services. Over several years, these parties worked to provide community programming (e.g., workshops, in-services). Eventually, community members requested a research project on services but reported they did not have the bandwidth to lead the project. Thus, all decided to engage in CBPR and work together in order to produce knowledge for advocacy (Wallerstein & Duran, 2006, 2010). There were no selection criteria other than interest from initial discussions. A next step was to tailor partnership.

#### Recognizing the Community as a Unit of Identity

Partners recognized the community as being grounded in race plus autism (Annamma et al., 2013; Crenshaw, 1989). While individuals have their own identities, whether one is perceived as minoritized shapes experiences (Hobbs, 2014). All partners shared experiences of racism, sexism, and ableism on the basis of how others perceived their skin tone, physiological traits, and differences from social norms. For example, one shared that others often assumed they were Latinx and spoke Spanish as a first language on the basis of their physical appearance; it did not matter that neither assumption corresponded to their actual identity. Others shared researchers were dismissive of them when they believed they were just minoritized and became inclusive when they learned they had lived experiences, but there was no one lived experience. Thus, partners defined the community as intersectional, recognizing heterogeneity was important.

#### Complex Contexts

In discussing the environment for CBPR, community partners felt researchers and other authority figures (e.g., doctors) used a deficit-driven lens to describe minoritized autistic individuals. Consistent with prior work norms (Onovbiona et al., 2023; Weitlauf et al., 2024), many felt practitioners assumed they were of low SES based on skin color or made assumptions about them on the basis of SES indicators (e.g., assuming they knew nothing about autism because of having a high school education). In both cases, SES failed to reflect their experiences (Powell, 2012; Swilley-Martinez et al., 2023). For example, while one community partner had a graduate degree and a secure job that provided for their family, being the sole caregiver for Black autistic children with constant support needs who faced racism in the educational system meant they were exhausted. Coupled with their own lived experiences as a Black adult with difficulty accessing language, this partner felt they had reduced access to services for themselves and their children. The partner felt the issue was not just racism or ableism, but rather, a confluence of factors. Given accounts like these, to show cultural humility and that the study was designed with minoritized autistic individuals in mind (Lewis & Oyserman, 2016), partners opted to focus on social-ecological factors. They felt including SES would require an investigation beyond the scope of this study, such as how U.S. immigration policy differently impacted diaspora grouped under one label (e.g., Latine) differed in ethnicity, professional background, and income.

#### Use of a Social-Ecological Model

Given multi-level domains of influence (Bronfenbrenner, 1977), partners chose to focus on topics that had spurred their decision to engage in CBPR: desired services and environmental impacts in their experiences (NIMHD, 2017). One community partner shared they and their spouse, who was also minoritized, struggled to obtain services for their child in secondary school. While they did not feel as sense of belonging to their community, given racism, they also felt school resistance to providing services was not due to racism. Rather, as a school-based provider, they believed this was typical behavior, as schools in their area were known for denying services in secondary education; in turn, they felt practitioners internalized this systems-wide practice. That is, practitioners reacted to the language skills of their child and decided services were not needed on the basis of community norms (not providing services), amid broader societal norms (federal policy for special education services).

#### Building Upon Community Strengths

In developing roles and procedures, given community strengths (CTSA, 2011), a focus was co-learning and deciding how to build capacity. In reviewing prior work (Nicolaidis & Raymaker, 2015), community partners found it did not meet their priorities. While race and disability were parts of them, they did not define them. Rather, partners felt their own identities and their communities were nuanced. Thus, instead of naming cultural values with a 1:1 ratio (e.g., respect is important in *x* culture), partners focused on sharing strengths and values in their community (e.g., learning about and keeping cultural knowledge and traditions alive across backgrounds). Next, partners related these strengths to one another and connected them to the research process, such the importance of lived experience for developing research methods and of community values for project completion (e.g., agency and cultural humility to support accessibility and collaboration). Building upon strengths also meant emphasizing academic partners could provide supports, but those did not replace community strengths.

#### Building Equitable Partnership

Developing equity in the partnership involved multiple levels of dynamics (Israel et al., 2003). Each partner recognized they had their own lived experiences, access needs, and life demands. Such humility facilitated flexibility in scheduling and co-learning. If a partner indicated written text was inaccessible and verbal communication was a strength, they received study materials in writing, discussion at meetings, and individual follow-up for further discussion. Relational humility also led to using consensus for decision-making. Each partner could propose a decision and share their opinion. Others were to listen to understand versus listen to respond. If consensus was not met, partners would re-enter discussion; however, this did not occur. Power sharing involved tailoring diversity representation and formal agreements. One partner relayed a white non-autistic researcher had invited them to join a community advisory board of “autistic people with one other identity.” While this reflected researcher bias (Major & O’Brien, 2005), it also led to tokenization (Maye et al., 2021). Other partners had similar experiences. Thus, all opted to have autonomy in how individuals represented themselves versus using positionality statements. Further, while partners developed a collective agreement on roles and responsibilities (e.g., research partners would consent participants, collect data, and present de-identified data; all would support conceptualization, study design, recruitment, analysis, and dissemination), formal agreements were not a cultural norm for all. Thus, partners regularly reviewed this agreement at meetings and considered it as a living document.

#### Balancing Knowledge with Action for Mutual Benefit

In planning research for action, community partners shared research priorities, which involved advocacy to inform policy (Israel et al., 2010). Research partners shared what they could support. Partners also discussed grants. In the past, researchers approached two community partners to “partner” on grants. One demanded a letter of support, which would have entailed the community partner completing a substantial amount of work, without asking if they wanted to be a partner or offering compensation. Another researcher paid minoritized autistic advisory board consultants $200 for five years of work but did not notify them that faculty consultants received over $200 per hour or that they had millions of dollars in grant funding. Here, the first author met with community partners to develop a plan for supporting them beyond the project (e.g., building professional skills) and grant proposals. In developing a proposal, community partners initially requested $50 per hour. When a faculty consultant later requested $200 per hour, the first author shared this information with partners, reviewed their contributions, asked if they wanted to adjust their rate, and revised the budget to $200 per hour for all consultants. Research partners also determined how to provide resources to community partners in non-monetary ways, such as by supporting organizations that aligned with community priorities (e.g., helping organizations obtain funding). In all cases, mutual benefit involved transparency in the research process.

#### Shared Community Dissemination

To disseminate findings, a key issue was balancing community partner involvement with equity (Israel et al., 2003). Community partners voiced priorities, which were to share findings locally and nationwide to increase awareness about services for self-advocates, families, and policymakers. This resulted in joint planning of an in-person community workshop, a virtual webinar for an advocacy network, a podcast, and social media posts. Community partners felt they had expertise in dissemination via social media and podcasting and asked for support in other areas. Accordingly, research partners coordinated logistics for the workshop and webinar, translated community partner ideas into preliminary dissemination materials, and revised them to align with community partner priorities. Research partners also took the lead on translating discussion about engagement strategies, scheduling, and event coordination (e.g., flow of activities, accessibility needs, set-up, personnel support) into action.

#### Commitment to Sustained Partnership for Change

The partnership informing this study began years before research, such that there was trust between community partners and the research team (Wallerstein et al., 2008). Yet once CBPR began, it was important for partners to directly discuss their commitment. Community partners spoke about their lived and family experiences with educational and healthcare systems. Their motivation was to have intersectional representation in the evidence base and to ensure CBPR led to advocacy and promotion of health equity. Research partners shared ways in which community partners were critical to science, clinical practice, and training (e.g., lack of representation means diagnostic criteria may fail to reflect the autistic population; Durkin et al., 2015). Their motivation for engaging in CBPR was to support communities by offering support (e.g., funding, personnel) as professionals with specific research skills that did not replace the expertise, experiences, priorities, or concerns of community partners. These interactions helped partners understand that researchers respected them for them, would advocate for them in different contexts (e.g., university, policymaking), and were committed to long-term partnership.

#### Engaging in Iterative, Cyclical CBPR

Engaging in CBPR resulted in new insight and research, with revision to the partnership. Community partners felt they had increased capacity to effect change, suggested new CBPR projects (e.g., self-determination in navigating care systems), and expressed an interest in taking on more power in research (e.g., as co-principal investigators and co-investigators). Research partners felt they had a better understanding of gaps in the evidence base and contexts underlying historical mistrust of research (Wallerstein et al., 2008). For example, a university wanted to feature partners in a donor publication. All partners agreed the coverage could support advocacy. However, after mutually agreeing upon photoshoot content, university administrators notified a junior research partner they changed the photoshoot without obtaining their consent. The photoshoot was to take place in another lab with equipment the partner did not use, as the university decided the lab and equipment of the research partner were not donor-worthy. This unilateral decision was of severe ethical concern to all, such that the research partner had to firmly explain to the university why they would not participate in the photoshoot. Ultimately, the university. This experience required reflection upon CBPR principles and underlined the responsibility of showing respect to communities through action (Wallerstein et al., 2019)

### Procedures

Participant selection criteria were: (a) racially minoritized, ethnically minoritized, or both racially and ethnically minoritized per U.S. Census guidelines, which included groups other than non-Hispanic white: American Indian or Alaska Native, Asian, Black or African American, Native Hawai’ian or Pacific Islander, multiracial, and other for race and Hispanic/Latine for ethnicity (Office of Management and Budget [OMB], 1997); (b) met DSM-5 criteria for autism (APA, 2013), per self-report of a formal diagnosis in screening and independent confirmation using the Social Responsiveness Scale-2^nd^ Ed. (SRS-2; Constantino, 2012) plus expert clinical judgment (i.e. administration by trained research staff and review by a clinical psychologist); (c) ages 13 to 30, coinciding with approximately when transition planning begins in the U.S. and 10 years post-federal eligibility for special education services (“Every Student Succeeds Act,” 2015); (d) proficiency in English per self-report during screening, as assessments were in English; (e) adequate hearing and vision thresholds for responding to human speech and computer screen images; and (f) use of primarily spoken language to communicate, as study activities required producing oral responses. Selection criteria for caregivers were: (a) active caregiving role for autistic participants; (b) proficiency in English, as interviews were completed in English; and (c) adequate hearing and vision thresholds for completing study activities. Exclusion criteria were: minimal spoken language, per self- or caregiver-report; and lack of adequate internet access. No participants were excluded, but if so, they were eligible to participate in a separate study on communication.

Recruitment took place in a multi-step process: (a) sharing study flyers with community and advocacy organizations serving minoritized autistic adolescents and adults by email, (b) providing consultation to interested individuals or individuals and families about the study in their modality of choice (phone, Zoom, email), (c) obtaining informed consent using a dynamic process, and (d) collecting data. The research team reviewed consent and assent forms, encouraged families to ask questions, and explained different options for sharing data (no data, de-identified data, transcripts, audio-recordings). Adults 18 and older who were their own legal guardian provided informed consent. Adults who were not their own legal guardians and those under 18 provided assent, and legal guardians provided informed consent. Recruitment and data collection took place from 2022 to 2023 remotely on HIPAA-compliant Zoom, using a stopping rule of *N* = 68 to account for possible 15% attrition. A Monte Carlo simulation determined the minimum detectable effect was 0.21 (a medium effect size), which was feasible for a sample of 60; however, participant referrals led to *N* = 73. The first author administered an assessment protocol, including questionnaires, to participants and caregivers, on Zoom.

### Measures

#### Race, Ethnicity, Sex at Birth, and Gender

Participants reported race, ethnicity, sex assigned at birth, and gender; see Table 1. Respondents selected one or more options for race: American Indian or Alaska Native, Asian, Black or African American, Native Hawai’ian or Pacific Islander, and white (OMB, 1997). Respondents could also write in options, which allowed for inclusion of categories added to the U.S. Census after data collection took place, such as Middle Eastern/North African (OMB, 2024). However, given few participants for specific combinations under multiracial, the team consulted the institutional review board office about privacy. If *n* < 5, full reporting was deemed to be potentially identifiable. Hence, reporting of multiracial categories is non-exclusive (i.e., someone in multiracial was also included in multiple racial categories, such as white and Black). Race, ethnicity, and gender were excluded from analysis, as they are complex constructs beyond the scope of this study (American Psychological Association, 2019). Sex at birth was also excluded, as prior work tends to confound sex at birth with gender (Halladay et al., 2015).

#### Language, NVIQ, and Autism Traits

Participants completed normed assessments across language domains (Girolamo et al., 2023b; Magiati et al., 2014), which align with clinical practice patterns for service eligibility in the U.S. (Burke et al., 2024; Selin et al., 2022); see Table 2. Overall receptive-expressive language was assessed by the Clinical Evaluation Language Fundamentals-5^th^ Ed., normed up to age 21 (*M* = 100, *SD* = 15; Wiig et al., 2013). Age 21 norms were used for participants over age 21, per prior work on adults ages 18 to 49 (Botting, 2020; Clegg et al., 2021; Fidler et al., 2011). Receptive and expressive vocabulary were assessed by the Peabody Picture Vocabulary Test-5^th^ Ed. (Dunn, 2019) and Expressive Vocabulary Test-3^rd^ Ed. (*M* = 100, *SD* = 15; Williams, 2019). Nonword repetition was assessed by percent accuracy on the Syllable Repetition Task (Shriberg et al., 2009). To contextualize language skills, given autism diagnostic criteria (APA, 2013; WHO, 2022), nonverbal general cognitive ability was assessed using the Raven’s Progressive Matrices-2^nd^ Ed. (*M* = 100, *SD* = 15; Raven et al., 2018). The Raven’s does not require verbal language for completion, which mitigates confounds in scores (Grondhuis et al., 2018). Last, given attention to the role of autism traits in patterns between language and NVIQ (Bal et al., 2016), autism traits were measured using the SRS-2, which has caregiver and self-report forms for students and adults (Constantino, 2012). Item scores provide subscale raw and *t*-scores for restricted and repetitive behaviors, social communication impairment, and overall autism traits (sub-clinical: 59 or below; mild: 60 to 65, moderate: 66 to 75; high: 76 or above).

#### Social-Ecological Factors

Sense of community was assessed by the Brief Sense of Community Scale, a validated scale for diverse youth and adults (Peterson et al., 2008). Respondents rate eight statements from strongly disagree to strongly agree on a five-point scale. Item scores are averaged to provide an overall score, with higher scores indicating higher sense of community. Current educational enrollment was assessed by respondent report: enrollment status in educational programming (yes/no) and type of programming if applicable (junior high, high school, specialized high school program, college courses, post-secondary program, other). To assess services, this study measured number of services received, unmet service needs, and barriers to services, as in Taylor and Henninger (2015). Respondents selected if they received each service: psychological or mental health; social work; speech-language therapy or communication; physical therapy; occupational therapy or life skills therapy or training; disability-related medical services; career counseling, job skills training, or vocational education; tutoring; transportation; assistive technology; audiology; orientation and mobility; reader or interpreter; respite care; and other. If services were not received, respondents indicated if each service was an unmet need and if each of 12 items were barriers to having service needs met; see Tables 3 and 4. Services received and unmet needs included “other,” with a write-in option.

### Data Processing

Two research assistants independently scored and checked measures. All disagreements were discussed until consensus was reached. NVIQ was auto-scored within its testing platform. Data were checked for missingness and replaced using predictive mean matching with one imputation in SPSS 29 (IBM Corp., 2023; Little & Rubin, 2019): language and NVIQ scores (*n* = 1; did not complete assessment); SRS-2 scores (*n* = 2; *n* = 1 missing form, *n* = 1 did not complete the SRS-2); and educational programming (*n* = 1; did not answer question). For the categorical approach, data were coded as follows: a) language impairment as ≤ −1.25 *SD* on ≥2 measures: CELF-5 Receptive Language Index, CELF-5 Expressive Language Index, SRT percent accuracy, PPVT-5 standard score, EVT-3 standard score (Girolamo & Rice, 2022; Tomblin et al., 1997); b) ID as NVIQ < 70 (APA, 2013); and high levels of autism traits as SRS-2 total *t*-scores > 76 (Constantino, 2012). For the continuous approach, language scores were checked for multicollinearity using variance inflation factors (VIF). As VIF ≥ 5, CELF-5 core language scores, which had the highest correlations with other language measures, were used. To facilitate interpretation, CELF-5 core language scores and NVIQ were centered on *M* = 100, and SRS-2 total *t*-scores were centered on the sub-clinical threshold of 59. Educational enrollment was coded as enrolled or not, as secondary and post-secondary distributions were similar and as Mann-Whitney U tests revealed nonsignificant differences in median counts for services received, *U* = 158, *z* = −.12, *p* = .916; unmet needs, *U* = 178, *z* = .49, *p* = .642; and barriers, *U* = 123.5, *z* = −1.18, *p* = .245.

### Analysis

To address the first research question, services questionnaires were analyzed descriptively to obtain means, standard deviations, and ranges for total number of services received, unmet service needs, and barriers to services. To address the second question, primary outcome measures were rate ratios and secondary outcomes were effect sizes. Analyses used an *a priori* significance level of .05. Given sample size, correlations identified significant effects to use in analysis. As data were non-normally distributed, Spearman (1904) correlations were used. Next, negative binomial regressions with a log link function and robust variance estimator separately estimated the extent to which number of services received, unmet service needs, and barriers to services were predicted from categorical traits (LI, ID, high level of autism traits) and social-ecological variables (sense of community, educational enrollment; Coxe et al., 2009). Negative binomial regression is appropriate for over-dispersed count data, in that the variance can be higher than the mean and the number of discrete events varies from person to person and setting to setting (Cameron & Trivedi, 2013; Gardner et al., 1995). Regression analyses were repeated with continuous traits (CELF-5 core language scores centered on 100, SRS-2 total *t*-scores centered on 59, and NVIQ centered on 100). Prior to analysis, model results were checked for linearity, normality, and homogeneity of variance.

## Results

### Participant Flow

Participants were 73 autistic individuals; see Table 1. Caregivers (*N* = 52) included 47 mothers, 2 grandparents, 2 fathers, and one sibling. The female-to-male ratio for sex assigned at birth and gender was approximately 1:2. Over half were enrolled in educational programming, 28 of whom were in secondary education. Most participants (54.79%) met criteria for LI; see Table 2. Six (8.22%) had NVIQ < 70 and met criteria for ID, and 15 (20.55%) had NVIQ of 70 to 84. Levels of autism traits varied: high (*n* = 33, or 45.21%), moderate (*n* = 19, or 26.03%), low (*n* = 10, or 13.70%), and subclinical (*n* = 11, or 15.1%).

### Patterns in Services Received, Unmet Service Needs, and Barriers to Services

On average, participants received more than three services (*M* = 3.67, *SD* = 2.76), with a range from 0 to 12; see Figure 1. Nearly all received at least one service (93.15%), over three-quarters received at least two services (76.71%), and more than one-half received at least three services (58.90%). A majority of participants received psychological or mental health services (65.75%) and disability-related medical services (54.79%), while approximately one-third received speech-language or communication services (34.25%); see Table 3. Participants reported over three unmet service needs (*M* = 3.32, *SD* = 3.20, range = 0-14). Over three-quarters reported at least one unmet service need (78.08%), and over two-thirds reported at least two service needs (67.12%). Of those not receiving a given service, the most common unmet service needs pertained to adulthood: career counseling or job skills training (64.71%) and occupational or life skills therapy or training (50%). The next highest unmet service need pertained to more global access, speech-language services and communication services (45.83%). Of services not received, most felt school-based services (e.g., in-class aide, tutor, reader or interpreter), access (assistive technology, audiology, orientation and mobility), and respite were not needed. Overall, even when receiving multiple services, participants had multiple unmet service needs.

**Figure 1.**
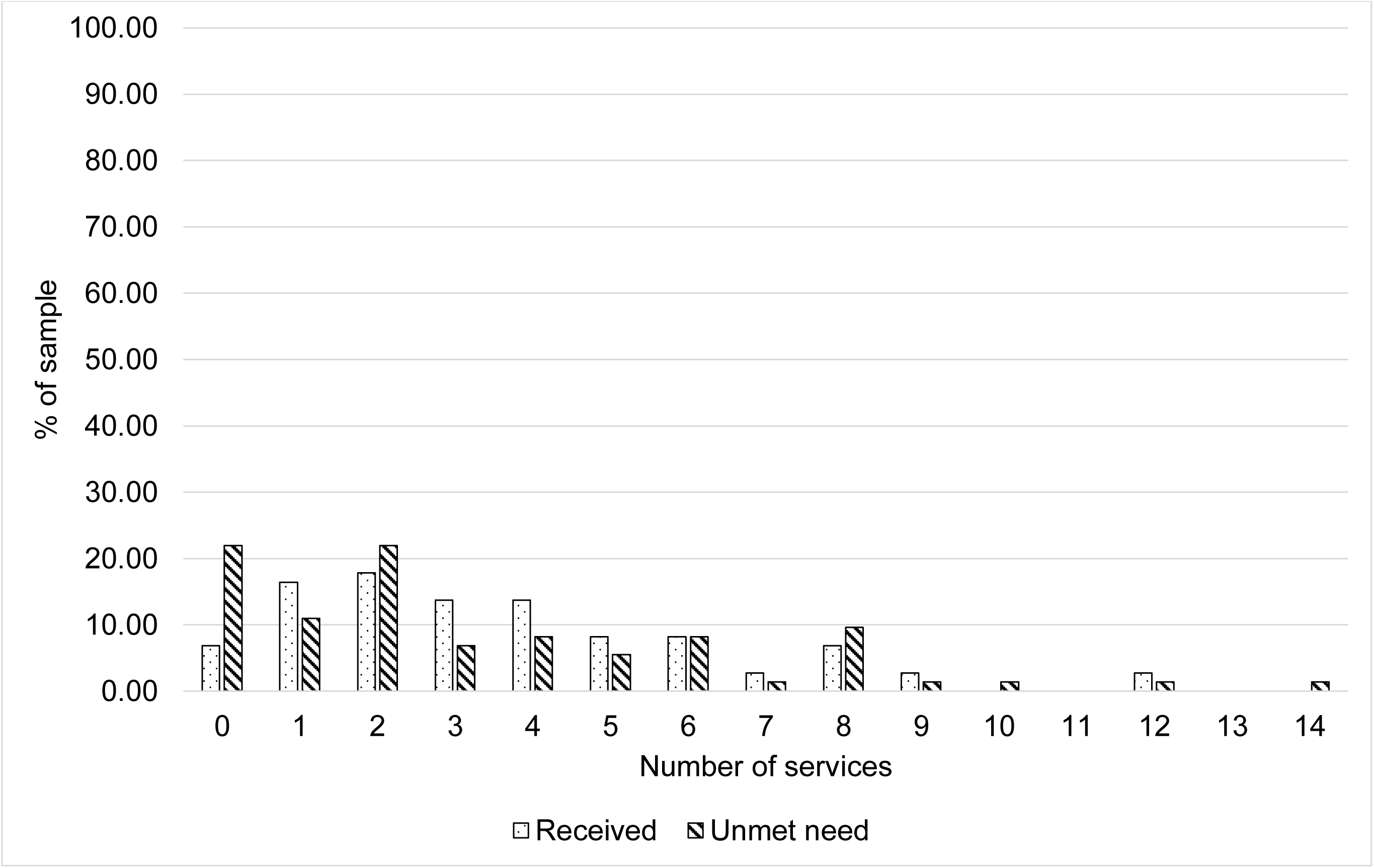
Frequencies of participants reporting the number of services received and unmet service needs

Participants also reported multiple barriers to services (*M* = 5.52, *SD* = 3.31, range = 0-11); see Table 4. A majority of the sample endorsed each of the 12 possible barriers to services, which primarily pertained to access: location (68.49%), services not available (67.12%), insurance not accepted (58.90%), ineligible (56.16%), cost (54.79%), getting information about services (54.79%), and scheduling conflicts (50.68%). Nearly all participants reported at least one barrier (87.67%), over three-quarters reported at least three barriers (78.08%), and over half reported at least six barriers (52.05%). In all, participants received multiple services, unmet needs, and barriers.

### Predictors of Services Received, Unmet Needs, and Barriers

#### Categorical Approach to Language, NVIQ, and Autism Traits

Spearman correlations revealed no significant relationships between services received, unmet needs, and barriers; see Table 5. Thus, receiving more services was not necessarily tied to fewer unmet needs or barriers. There were small to moderate effects of individual difference and social-ecological variables on each services outcome. Receiving more services was associated with having LI, having ID, and being enrolled in education. Descriptively, effect sizes of LI and educational enrollment were nearly twice that of ID. High levels of autism traits and lower sense of community were associated with more unmet service needs, and to a lesser extent, more barriers. There were few patterns among predictors. Having LI was associated with having ID, and lower sense of community was associated with high levels of autism traits.

Separate negative binomial regression models significantly predicted the number of services received and unmet service needs but not barriers; see Table 6. In the first model, for an individual without ID, without LI, and not in educational programming, the number of services received was 1.97. Holding all other variables constant, the number of services received was 1.70 times greater if an individual had LI than not (4.42 versus 2.60 services) and 1.56 times greater if in educational programming versus not (4.24 versus 2.71 services). The effect of ID was not significant. Next, analysis examined unmet service needs. The number of unmet service needs was 5.52 for an individual without high levels of autism traits and a sense of community score of 3.06, which roughly corresponds with neither agree nor disagree. Accounting for all variables, the number of unmet needs in individuals with high levels of autism traits was 1.82 times greater than those without (i.e., lower than) high levels of autism traits (4.29 versus 2.36 needs). In sum, receiving more services was tied to LI and educational enrollment, while unmet service needs only involved high levels of autism traits.

#### Continuous Approach to Language, NVIQ and Autism Traits

When using continuous measures of overall language, NVIQ, and autism traits, effects somewhat differed from the categorical approach; see Table 7. Services variables did not necessarily correspond with one another, but there was a significant, moderate positive association between barriers to services and unmet service needs. As in the categorical approach, there were small to moderate effects of language, NVIQ, and educational enrollment on services received. Lower language scores, lower NVIQ, and being in educational programming significantly associated with receiving more services, but the effect size of NVIQ was 1.75 times larger than that of intellectual disability (ρ = −.42 versus ρ = .24). Effects of SRS-total *t*-scores and sense of community on number of unmet service needs and barriers were similar to the categorical approach. Higher SRS-2 total *t*-scores associated with more unmet needs and barriers, and sense of community showed the opposite pattern. Other differences pertained to predictor variables. Lower sense of community associated with higher SRS-2 total *t*-scores, and higher CELF-5 core language scores associated with NVIQ. However, the effect size of NVIQ with CELF-5 scores was over two times larger than that of intellectual disability (ρ = .59 versus ρ = .27). Last, higher CELF-5 core language scores were associated with not being in educational programming. In all, patterns varied when using a continuous approach to language, NVIQ and autism traits: 1) significant associations between unmet needs and barriers, as well as CELF-5 core language scores and educational enrollment, and 2) stronger associations between NVIQ than intellectual disability and CELF-5 core language scores, as well as educational enrollment.

As in the categorical approach, separate negative binomial regression models were statistically significant for number of services received and unmet service needs but not barriers; see Table 8. However, effects differed. Number of services received was predicted by educational programming, but not CELF-5 core language scores or NVIQ. For a participant with NVIQ of 100, CELF-5 core language score of 100, and not in educational programming, the average expected number of services received was 2.11. Holding all other variables constant, the number of services received was 1.48 times greater if in educational programming versus not (4.07 versus 2.75 services); in comparison, the rate ratio was 1.56 in the categorical approach. Turning to unmet service needs, for an individual with a sense of community score of 3.06 (roughly indicating neither agree nor disagree) and an SRS total *t*-score of 59 (corresponding to the sub-clinical threshold for autism traits), the expected number of unmet service needs was 4.50. When accounting for all other variables, the number of unmet needs was 1.03 times greater for every one-unit increase in SRS-2 total *t*-scores above 59; in contrast, the rate ratio of high level of autism traits was 1.82. Findings indicate receiving more services was tied only to educational enrollment, while unmet service needs appeared to be tied to SRS-2 total *t*-scores.

### Summary

Participants received multiple services yet reported multiple unmet service needs and barriers to having service needs met. Categorical versus continuous approaches to characterizing language, NVIQ, and autism traits yielded different outcomes in correlation and analyses. Some correlations became nonsignificant in regression. In particular, LI, but not language scores, predicted receiving more services.

## Discussion

In evaluating services in minoritized autistic adolescents and adults varying in language skills, this study used CBPR to focus on the intersection of race and disability. All partners agreed upon conceptual frameworks for contexts underlying CBPR and services (Annamma et al., 2013; Plaut, 2010). This approach informed interpretation of the findings.

### Clinical Implications

The premise of this study was that broad inclusion in research is crucial for evidence-based practice (Sackett et al., 1996). Participants, 39.73% of whom were in high school, reported 3.34 services received and 3.32 unmet needs. Compared to prior work, the number of services received was similar to autistic youth and adults in their last year of high school (*n* = 3.05), while the number of unmet needs was higher (*n* = 1.85; Taylor & Henninger, 2015). In contrast, the number of services received was lower than autistic adolescents and adults (*n* = 6.13), 40.8% of whom were in high school, while the number of unmet needs was similar (*n* = 3.18; Ishler et al., 2022, 2023). Beyond high school enrollment and racial diversity, studies differed in other ways. Less than 10% of participants had co-occurring ID using an NVIQ measure versus 27% via caregiver report in Ishler et al. (2022) and 30.8% in Taylor and Henninger (2015) using an IQ measure that requires verbal language. Without directly comparable measures, determining similarities across samples is challenging. Regardless, individual service variables suggest there were differences. Relative to other studies, participants in this study were approximately two-thirds to over two times less likely to receive transportation or social work services (Ishler et al., 2023); three to eight times more likely to endorse speech-language, transportation, and personal assistant or aide services as unmet service needs (Taylor & Henninger, 2015); and more likely to endorse each barrier to services except for language (Koffer Miller et al., 2022; Newman et al., 2011; Taylor & Henninger, 2015); see Supplementary Table 3. While mean counts for services received and unmet needs appear similar, closer inspection points to qualitative differences.

A second dimension of nuance involves different approaches to measuring clinical characteristics. Care systems often use categorical cutoffs, such as LI or ID (ESSA, 2015; IDEIA of 2004, 2018), despite heterogeneity in both language and NVIQ across the autism spectrum (Bal et al., 2016; Schaeffer et al., 2023). Here, a continuous approach (language scores, NVIQ, autism trait scores) yielded different patterns than a categorical approach (LI, ID, high level of traits): there were significant associations between language and educational enrollment which were nonsignificant in the categorical approach, and there was a stronger association between language and NVIQ than in the categorical approach. These differences raise the question of clinical significance. For instance, Johnson et al. (1999) found a research definition of LI in autistic and nonautistic adults yielded a higher estimate than clinical judgment of LI (11.7% versus 5.5%). Here, over 55% of participants qualified for LI (Tomblin et al., 1997), which predicted the number of services received. Nearly two-thirds (64.39%), however, received speech-language services (34.25%) or reported speech-language services as an unmet need (30.14%). Given this disproportionality, it is unknown whether the operational definition of LI in this study aligns with clinical judgment or if aspects of language (e.g., pragmatics, discourse) influenced service patterns (Schaeffer et al., 2023). Ultimately, differences across samples and approaches to measurement underline the importance of assessing all areas where individuals may want support (Musgrove, 2015).

### Implications for Autistic Adolescents and Adults

Findings also point to the relevance of contextual factors in services (Bronfenbrenner, 1977; NIMHD, 2017). Predictors did not significantly predict barriers to services. However, higher levels of autism traits and a lower sense of community were each associated with one another, as well as with more barriers. Taking race and disability as social constructs (Annamma et al., 2013), it could be that barriers to services, sense of community, and autism traits reinforce one another. On one hand, minoritized autistic individuals and families face systemic barriers to services, including lack of practitioner knowledge about cultural norms (Onovbiona et al., 2023; Pham & Charles, 2023; Weitlauf et al., 2024), amid little knowledge overall on sociocultural variation in perceptions about autism (Golson et al., 2022). Beyond hindering access to services, these gaps may negatively impact sense of community (Gulsrud et al., 2021; Michalski et al., 2020). Barriers to services may also explain why minoritized autistic youth are disproportionately likely to have high unmet support needs (Hughes et al., 2023). Elucidating relationships between barriers to services, contextual factors (e.g., sense of community), and clinical expertise (Wolfson et al., 2000) using person-centered measures is critical for developing evidence-based practice to promote health equity (Sackett et al., 1996).

This study also tailored CBPR for a localized context, given a lack of precedent for complex contexts that community partners saw as relevant, such as disclosure (Fletcher-Watson et al., 2019), individual identity (Nicolaidis & Raymaker, 2015), and equity in partnership, mutual benefit, and community dissemination (Israel et al., 2003). To share power (Wallerstein et al., 2019), partners used the same approach to conceptualize representation on the team as in participants: attending to heterogeneity within minoritized autistic individuals versus checking a box (Plaut, 2010). This approach provided a pathway to partnering with community members who research systematically excludes as members of the scientific community and as participants (Maye et al., 2021). Sustaining these inclusive partnerships is the next step to producing the best-available evidence and clinical practice (Sackett et al., 1996).

### Limitations

This study encountered several limitations. First, analysis did not include SES, which has shown inconsistent effects on services (Eilenberg et al., 2019; Levine et al., 2007). As a complex construct (American Psychological Association, 2021), prior effects of SES were unclear. Some studies examined household income and found negative effects on services received and unmet service needs (Laxman et al., 2019; Shattuck et al., 2011) or no effects on services received, unmet service needs, and barrier (Ishler et al., 2023; Taylor & Henninger, 2015). Family financial burden, which is specific to individual need, predicted barriers to services (Ishler et al., 2023); however, it may not be broadly applicable, such as for individuals who do not have families. This study also did not examine associations between services and age. While federal special education affords eligibility for services until age 21 (“IDEIA of 2004,” 2018), compulsory special education age limits vary from 18 to 22 (National Center for Education Statistics, 2024) and rose during the pandemic. Without detailed information on individual eligibility, age and services associations may be challenging to interpret. A second limitation was not reporting detailed participant information, including exact multiracial categories or type of educational programming. Concerns about making participants re-identifiable (O’Keefe & Rubin, 2015), plus institutional review board consultation, led to not reporting precise information. Larger sample sizes are needed to balance privacy with full reporting. Third, this study used counts to measure services. As supported by qualitative evidence (Anderson et al., 2018a), more nuanced measures, such as self-ratings of the extent of unmet service needs (Burke et al., 2024), may better inform our understanding of services.

### Future Directions

Findings and limitations chart pathways forward to advance our understanding of services in minoritized autistic adolescents and adults for advocacy. First, heterogeneity within a 100% minoritized sample in the presence and nature of services justified use of DisCrit and Diversity Science (Annamma et al., 2013; Plaut, 2010). What is unknown is how these theories inform larger-scale modeling of the transition to adulthood when accounting for both nuances in individual language and cognitive profiles plus environmental factors in services access. Prior work has examined broad patterns in large datasets (Song et al., 2022; Turcotte et al., 2016) and longitudinal change in individual studies (Laxman et al., 2019), but future work is needed to merge theory-founded approaches to heterogeneity with statistical methods. A second area for future research is improved measures for services (Burke et al., 2024), as well as social-ecological correlates (Anderson et al., 2018b). Here, sense of community was associated with services, but the clinical significance of this relationship is unknown. Given sociocultural variation in perceptions about autism (Rivera-Figueroa et al., 2022), understanding what sense of community in the context of services means across different individuals is complex. Determining how to implement and use self-rating measures for services and sense of community may require particular attention and care (Douglas et al., 2022). These directions for research serve to advance knowledge and advocacy for minoritized autistic adolescents and adults.

## Conclusion

In using CBPR to study services received, unmet needs, and barriers in minoritized autistic adolescents and adults, this study began with lived experiences as expert opinion to guide investigation. Using theory-driven approaches to investigation, findings showed heterogeneity in the presence and nature of services. Correlates of services differed when using continuous or categorical approaches to language, NVIQ, and autism traits, further supporting the relevance of heterogeneity in characterizing the transition to adulthood. This approach and findings may support iterative development of evidence-based practice.

## Data Availability

All data produced in the present study are unavailable due to participants opting not to share their de-identified data in a dynamic consent process.

## Acknowledgements

We thank participants, families, community partners, and student assistants. This work was supported by an American Speech-Language-Hearing Foundation New Investigators Research Grant (PI: Girolamo) and NIH L70DC021323 (PI: Girolamo).

## Acknowledgements

TG was supported by an American Speech-Language-Hearing Foundation New Investigators Research Grant (PI: Girolamo) and NIH L70DC021323.

