## Supplemental Tables for "Services in Minoritized Autistic Adolescents and Adults Varying in Language Skills"

| **Supplementary Table 1**  *Services Received and Unmet Service Needs in Prior Studies of Autistic Adolescents and Adults* | | | | | | | | | | | | | | |
| --- | --- | --- | --- | --- | --- | --- | --- | --- | --- | --- | --- | --- | --- | --- |
| **Service** | Ishler et al. (2022, 2023) | | NLTS2 | | Shattuck et al. (2011) | | Song et al. (2022) | | Taylor & Henninger (2015) | | Turcotte et al. (2016) | | | |
|  |  |  |  |  |  |  |  |  |  |  | M/HS | | Adult | |
|  | Rec | Unmet | Rec | Unmet | Rec | Unmet | Rec | Unmet | Rec | Unmet | Rec | Unmet | Rec | Unmet |
| **Personal counseling** |  |  |  |  |  |  |  |  |  |  |  |  |  |  |
| 1. Psychological/mental health services or counseling | 62.6 | NR | 46.2 | - | 35 | - | 38-40 | 19-21 | 43.6 | 15.4 | 47.9 | 13.8 | 46.8 | 17.3 |
| 2. Social work service | 53.4 | NR | 27.7 | - | 41.9 | - | 38-48 | 17-22 | 15.4 | 15.4 | 59.6 | 8.5 | 63.5 | 11.2 |
| **Therapeutic services** |  |  |  |  |  |  |  |  |  |  |  |  |  |  |
| 3. Speech-language therapy/communication services | 35.6 | NR | 74.6 | - | 9.1 | - | 10-40 | 7-21 | 35.9 | 10.3 | 54.2 | 7.9 | 9.9 | 21.9 |
| 4. Physical therapy | - | - | 17.3 | - | - | - | 10-17 | 10-16 | 5.1 | 0 | 13 | 9.1 | 4.8 | 8.2 |
| 5. Occupation/life skills therapy or training | 44.8 | 33.3 | 49 | - | - | - | 12-31 | 14-22 | 25.6 | 30.8 | 39 | 16.5 | 12 | 21.2 |
| **Health-related services** |  |  |  |  |  |  |  |  |  |  |  |  |  |  |
| 6. Disability-related medical services | 65.5 | NR | 46.9 | - | 23.5 | - | 18-34 | 8-14 | 30.8 | 10.3 | 19.2-60* | 3.3-10.3 | 24.3-63.6* | 5.8-11.8 |
| **Vocational services** |  |  |  |  |  |  |  |  |  |  |  |  |  |  |
| 7. Career counseling or vocational/job skills training | 54 | 33.3 | 20.6 | - | - | - | - | - | 33.3 | 35.9 | - | - | - | - |
| **Academic enhancements** |  |  |  |  |  |  |  |  |  |  |  |  |  |  |
| 8. Tutoring | - | - | 14.3 | - | - | - | - | - | 25.6 | 15.4 | - | - | - | - |
| **Services to increase access and mobility** |  |  |  |  |  |  |  |  |  |  |  |  |  |  |
| 9. Transportation services | 44.3 | - | 54.6 | - | - | - | - | - | 20.5 | 7.7 | - | - | - | - |
| 10. Assistive technology services/devices | - | - | 15.7 | - | - | - | - | - | 10.3 | 10.3 | - | - | - | - |
| 11. Audiology services for hearing problems | - | - | 2.5 | - | - | - | - | - | 0 | 7.7 | - | - | - | - |
| 12. Orientation and mobility services | - | - | 4.8 | - | - | - | - | - | 0 | 2.6 | - | - | - | - |
| **Personal assistance** |  |  |  |  |  |  |  |  |  |  |  |  |  |  |
| 13. Reader or interpreter | - | - | 6.1 | - | - | - | - | - | 10.3 | 0 | - | - | - | - |
| 14. Respite care | - | - | 19.6 | - | - | - | - | - | 10.3 | 5.1 | - | - | - | - |
| 15. Personal assistant or in-home/in-classroom aide | - | - | 53.9 | - | - | - | 38-46 | 19-24 | 33.3 | 2.9 | 42.7 | 13.1 | 24.9 | 21 |
| **Other** |  |  |  |  |  |  |  |  |  |  |  |  |  |  |
| 16. Other services | - | - | 4.7 | - | - | - | - | 10-16 | 5.1 | 15.4 | - | - | - | - |
| *Note.* M/HS = Middle or high school. Rec = service received. Unmet = unmet service need. NLTS2 = National Longitudinal Transition Survey 2 (United States Department of Education, National Center for Special Education Research, 2014). - = study did not assess. NR = not reported. M/HS = middle/high school. Song et al. (2022) reported as range for adolescence, transition-age adults, and young adults.  *Lower bound = neurology services. Upper bound = medication management. | | | | | | | | | | | | | | |

| **Supplementary Table 2**  *U.S.-Based Studies Examining Services, Unmet Service Needs & Barriers to Services in Autistic Adolescents and Adults* | | | | | | | |
| --- | --- | --- | --- | --- | --- | --- | --- |
| Author | Participant | Autistic Individual Characteristics | | | | | Services  (*M* ,*SD*, range) |
|  |  | *N* | Age | Race/Ethnicity | Sex/Gender | Co-Occur |  |
| Ishler et al. (2022) | caregivers | 168 | 16-30 | **Black, Hispanic, multiracial:** 21.3%  **Not:** 78.7% | **Gender**  28.2% female  71.8% male | **ID:** 27%  **Mental health:** 60.9%  **Medical:** 23.6% | **R:** 6.13 (3.23), 0-15 of 15  **U:** 3.18 (2.56), 0-11 of 15 |
| Ishler et al. (2023) |  |  |  | **Black, Hispanic, Native American or Alaska Native, multiracial:** 20.7%  **Not:** 79.3% |  |  |  |
| Laxman et al. (2019) | caregivers | 204 | 10.8-23.5 | **white:** 93.6%  **other:** 6.4% | **Sex/Gender**  25.5%/27% female  74.5%/70% male  3% other gender | **ID:** 58.8% | **R:** NR of 9  **U:** NR of 9 |
| Koffer Miller et al. (2022) | self | 1204 | 27.8 | **white:** 82.6%  **others:** 17.4% | **Sex**  28.2% female  71.8% male | **ID:** 21.5%  **Mental health:** 77% | **R:** NR of 21  **U:** NR of 21  **B:** NR of 8 |
| Shattuck et al. (2011) | caregivers | 410 | 19-23 | **white:** 74.8%  **Black:** 16.4%  **other or mixed:** 8.7% | **NR**  14.2% female  85.8% male | **ADHD:** 34.5%  **Severe LI:** 21.2% | **R:** 39.1% 0 of 4 |
| Taylor & Henninger (2015) | caregivers | 39 | 17-22 | **white:** 89.7%  **other race/ethnicity:** NR | **NR**  79.5% male | I**D:** 30.8%  **Psychiatric:** 51.3% | **R:** 3.05 (2.46), 0-9 of 16  **U:** 1.85 (1.94, 0-7) of 16 |
| Song et al. (2022) | caregivers | 3527 | 12-31+ | **white:** 77%  **Black:** 7%  **other:** 16% | **Gender**  21% female  79% male | **ID:** 17%  **Mental health:** 58% | **R:** NR of 9  **U:** NR of 9 |
| Turcotte et al. (2016) | caregivers | 1842 | 15.1-25.3 | **white:** 92.8%  **Black:** 7.2% | **Gender**  18.9% female  81.3% male | **ADHD:** 36%  **DD:** 35%  **ID:** 21%  **LD:** 27% | **R:** NR of 9  **U:** NR of 9 |
| *Note.* Co-occur = co-occurring diagnosis. White = non-Hispanic white. ID = intellectual disability. Mental health = mental health diagnosis. Medical = severe medical condition. R = services received. U = unmet service need. B = barrier to having service needs met. NR = assessed and not reported. - = not assessed. LI = language impairment. ADHD = attention deficit hyperactivity disorder. Services received and unmet needs list total possible maximum. Burke & Keller (2017) excluded, as no separate analyses are presented for autism. DD = developmental delay. LD = learning disability. Song et al. (2022) and Turcotte et al. (2016) analyzed the same dataset, but Turcotte et al. (2016) includes middle/high school and adults, and Song et al. (2022) included adolescents, transition-aged adults, and older adults. Turcotte et al. (2016) reported but Asian/Pacific Islander, Latino, Hispanic, or Chicano, Native American participants but excluded them from analysis. Ishler et al. (2022) defined under-represented minority race/ethnicity as Black, Hispanic, multiracial, reporting caregiver demographics. Ishler et al. (2023) was based on the same sample and defined “historically marginalized minority race/ethnicity” as Black, Hispanic, Native American/Alaska Native, or multiracial, reporting autistic individual demographics. | | | | | | | |

| **Supplementary Table 3**  *Comparison of Barriers to Having Service Needs Met in Present Sample (N = 73) with Prior Studies of Autistic Adolescents and Adults* | | | | | |
| --- | --- | --- | --- | --- | --- |
| **Barrier** | Full Sample | Ishler et al. (2023)* | Koffer Miller et al. (2022) | NLTS2 | Taylor & Henninger (2015) |
| 1. Location of services | 68.49 | 1.11 (1.15) | 15.5-19.6** | 32.7 | 33.3 |
| 2. Services not available | 67.12 | 1.32 (1.32) | 15.5-19.6** | 49.3 | 25.6 |
| 3. Doctor/specialist does not accept insurance | 58.90 | 0.73 (1.11) | 15.3*** | - | 33.3 |
| 4. Ineligible for services | 56.16 | 1.01 (1.25) | 3.2-6.4**** | 33.2 | 23.1 |
| 5. Cost of services | 54.79 | 1.03 (1.22) | 15.3*** | 33.9 | 38.5 |
| 6. Getting information about services | 54.79 | 1.56 (1.20) | - | 20.5 | 40.4 |
| 7. Scheduling conflicts | 50.68 | 0.91 (1.08) | 12.5 | 29.5 | 23.1 |
| 8. Lack of time for services | 42.47 | 1.03 (1.05) | - | 26.3 | 12.8 |
| 9. Poor service quality | 42.47 | 1.05 (1.21) | - | 34.6 | 15.4 |
| 10. Transportation | 41.10 | 0.81 (1.14) | 16.7 | 18.9 | 17.9 |
| 11. Physical accessibility | 6.85 | 0.23 (0.72) | - | - | 2.6 |
| 12. Language barrier | 1.37 | 0.45 (0.90) | - | 14.3 | 2.6 |
| *Note.* Barriers organized by frequency. For barriers with identical frequencies, list is in alphabetical order. - = study did not assess. NR = not reported.  *Scores reported on a scale of 0-3 as *M* (SD).  **Original items = no service providers in the area (15.5%) and not enough service providers in the area (19.6%).  ***Items were collapsed in Miller et al. (2022).  ****Original items = providers in the area will not see people with autism & providers in the area will not see people with mental health diagnosis. | | | | | |
