## Supplementary material for "Services in Minoritized Autistic Adolescents and Adults Varying in Language Skills": Tables

| **Table 1**  *Participant Sociodemographics* | | |
| --- | --- | --- |
| Variable | All (*N* = 73) | |
|  | *n* | *%* |
| Chronological age | 19.69 (4.71), 13.27-30.43 | |
| Race |  |  |
| American Indian, Native American, Alaska Native: alone | 5 | 6.85 |
| Asian | 8 | 10.96 |
| Black: alone | 29 | 39.73 |
| multiracial | 26 | 35.62 |
| Pacific Islander: alone | 4 | 5.48 |
| White: alone | 3 | 6.4 |
| Other | 3 | 4.11 |
| Hispanic/Latine | 13 | 17.81 |
| Sex Assigned at Birth |  |  |
| Female | 23 | 31.51 |
| Male | 50 | 68.49 |
| Gender |  |  |
| Female | 24 | 32.88 |
| Male | 49 | 67.12 |
| In-school status: Yes | 42 | 55.16 |
| Secondary education | 29 | 39.73 |
| College courses | 4 | 5.48 |
| Post-secondary program | 9 | 12.33 |
| In-school status: No | 31 | 42.47 |
| *Note.* Age presented as *M* (*SD*), range. Other included: Puerto Rican (*n*=2), Latine (*n*=2). Exact multiracial categories per participant are not reported to uphold participant privacy and confidentiality. If considering participants who reported more than one racial category, sample frequencies for race are: *n* = 19 (26.03 %) for Asian, *n* = 38 (52.05%) for Black, *n* = 4 for Pacific Islander (5.48%), *n* = 20 (27.40%) for white, and *n* = 9 (12.33%) for other. Other included: *n* = 1 (1.37%) for Middle Eastern, *n* = 3 (4.11%) for Latine, *n* = 2 (2.74%) for Puerto Rican, and *n* = 3 (4.11%) not reported. | | |

| **Table 2**  *Participant Language, NVIQ, and Autism Traits* | | | |
| --- | --- | --- | --- |
| Variable | Full Sample (*N =* 73) | | |
|  | *M* | *SD* | range |
| CELF-5 Receptive Language Index standard score | 82.06 | 20.39 | 45-118 |
| CELF-5 Expressive Language Index standard score | 77.40 | 18.84 | 45-116 |
| PPVT-5 standard score | 92.03 | 20.58 | 40-129 |
| EVT-3 standard score | 93.97 | 17.71 | 49-128 |
| Syllable Repetition Task percent accuracy | 90.50 | 9.22 | 66-100 |
| Raven's 2 NVIQ standard score | 90.68 | 16.14 | 47-135 |
| SRS-2 total *t*-score | 72.65 | 11.40 | 51-90 |
| Brief Sense of Community Scale average score | 3.06 | 0.86 | 1.38-5 |
| *Note.* CELF-5 = Clinical Evaluation of Language Fundamentals-5^th^ Ed. (Wiig et al., 2013). Peabody Picture Vocabulary Test-5^th^ Ed. (Dunn, 2019). Expressive Vocabulary Test-3^rd^ Ed. (Williams, 2019). Syllable Repetition Task (Shriberg et al., 2009). Raven’s 2 = Raven’s 2 Progressive Matrices (Raven, 2018). SRS-2 = Social Responsiveness Scale-2^nd^ Ed. (Constantino, 2012). NVIQ = nonverbal intelligence. Scores replaced using single imputation of variable means for: CELF-5, PPVT-5, EVT-3, SRT, and NVIQ scores (*n* = 1), as well as SRS-2 total *t*-scores (*n* = 2). Twenty-five participants were ages 22 or older. Brief Sense of Community Scale, with a range of 0 to 5, or strongly disagree to agree (Peterson et al., 2008). | | | |

| **Table 3**  *Frequencies of Services Received and Unmet Service Needs (N = 73)* | | | |
| --- | --- | --- | --- |
| Service |  | | |
|  | Receiving (%) | Not Received | |
|  |  | Unmet Need (%) | Not Needed (%) |
| 1. Psychological/mental health services or counseling | 65.75 | 15.07 | 19.18 |
| 2. Medical services or diagnosis/evaluation related to  special needs | 54.79 | 12.33 | 32.88 |
| 3. Speech-language therapy/communication services | 34.25 | 30.14 | 35.62 |
| 4. Occupation/life skills therapy or training | 31.51 | 34.25 | 34.25 |
| 5. Career counseling or vocational/job skills training | 30.14 | 45.21 | 24.66 |
| 6. Transportation services | 27.40 | 30.14 | 42.47 |
| 7. Personal assistant or in-home/in-classroom aide | 21.92 | 23.29 | 54.79 |
| 8. Social work service | 21.92 | 28.77 | 49.32 |
| 9. Tutor | 20.55 | 27.40 | 52.05 |
| 10. Assistive technology services/devices | 16.44 | 15.07 | 68.49 |
| 11. Other services | 16.44 | 26.03 | 57.53 |
| 12. Respite care | 16.44 | 23.29 | 60.27 |
| 13. Physical therapy | 12.33 | 10.96 | 76.71 |
| 14. Audiology services | 8.22 | 8.22 | 83.56 |
| 15. Orientation and mobility services | 2.74 | 9.59 | 87.67 |
| 16. Reader or interpreter | 2.74 | 16.44 | 80.82 |
| *Note.* Services organized by how frequently respondents selected "yes" to receipt of service. Services with identical frequencies are presented in alphabetical order. Received and not received services reflect frequencies in the entire sample. | | | |

| **Table 4**  *Barriers to Having Service Needs Met (N = 73)* | |
| --- | --- |
| Service | Full Sample  (*N* = 73) |
|  | % |
| 1. Location of services | 68.49 |
| 2. Services not available | 67.12 |
| 3. Doctor/specialist does not accept insurance | 58.90 |
| 4. Ineligible for services | 56.16 |
| 5. Cost of services | 54.79 |
| 6. Getting information about services | 54.79 |
| 7. Scheduling conflicts | 50.68 |
| 8. Lack of time for services | 42.47 |
| 9. Poor service quality | 42.47 |
| 10. Transportation | 41.10 |
| 11. Physical accessibility | 6.85 |
| 12. Language barrier | 1.37 |
| *Note.* Barriers organized by frequency selected. Barriers with identical frequencies are presented in alphabetical order. | |

| **Table 5**  *Spearman's Correlations Coefficients of Services, Categorical Individual Differences, and Social-Ecological Factors* | | | | | | | | |
| --- | --- | --- | --- | --- | --- | --- | --- | --- |
|  | 1 | 2 | 3 | 4 | 5 | 6 | 7 | 8 |
| 1. Number of services | - |  |  |  |  |  |  |  |
| 2. Unmet service needs | -.00 | - |  |  |  |  |  |  |
| 3. Barriers to services | .17 | .17 | - |  |  |  |  |  |
| 4. Language impairment: Yes | .40** | .05 | .16 | - |  |  |  |  |
| 5. Intellectual disability: Yes | .24* | .03 | .13 | .27* | - |  |  |  |
| 6. Autism traits: High | -.06 | .42** | .25* | -.14 | -.05 | - |  |  |
| 7. Sense of community | .16 | -.44** | -.27* | .04 | -.16 | -.38** | - |  |
| 8. Educational enrollment: Yes | .39** | -.02 | .11 | .20 | .16 | -.22 | .11 | - |
| *Note.* Barriers to services = barriers to having services needs met. Sex = sex assigned at birth.  * *p* < .05. ** *p* < .01. Intellectual disability and services received *p* = .045. Autism traits and barriers *p* = .028. | | | | | | | | |

| **Table 6**  *Negative Binomial Regression Results of Numbers of Services, Unmet Needs, and Barriers to Having Service Needs Met Using a Categorical Approach to Individual Differences* | | | | | |
| --- | --- | --- | --- | --- | --- |
|  | RR | 95% CI | *χ^2^* | *df* | *p* |
| *Number of Services* | | | | | |
| Model |  |  | 20.70 | 3 | < .001 |
| Intercept | 1.97** | 1.39, 2.81 |  |  |  |
| Intellectual disability: Yes | 1.11 | .78, 1.58 |  |  |  |
| Language impairment: Yes | 1.70** | 1.20, 2.39 |  |  |  |
| Educational enrollment: Yes | 1.56* | 1.11, 2.21 |  |  |  |
| *Unmet Service Needs* | | | | | |
| Model |  |  | 15.37 | 2 | < .001 |
| Intercept | 5.52** | 2.19, 13.94 |  |  |  |
| High level of autism traits: Yes | 1.82* | 1.14, 2.91 |  |  |  |
| Sense of community | .76 | .57, 1.01 |  |  |  |
| *Barriers to Services* | | | | | |
| Model |  |  | 1.99 | 2 | .371 |
| Intercept | 8.71 | 5.85, 12.99 |  |  |  |
| High level of autism traits: Yes | 0.79 | .59, 1.06 |  |  |  |
| Sense of community | 0.90 | .77, 1.05 |  |  |  |
| *Note.* RR = rate ratio. All significant rate ratios at *p* < .05 are bolded. Intellectual disability = NVIQ < 70. Language impairment = ≤ -1.25 *SD* on ≥ 2 language measures: Clinical Evaluation of Language Fundamentals-5 (CELF-5) Receptive Language Index, CELF-5 Expressive Language Index (Wiig et al., 2013), Syllable Repetition Task percent accuracy (Shriberg et al., 2009), Peabody Picture Vocabulary Test-5 standard score (Dunn et al., 2019), and Expressive Vocabulary Test-3 standard score (Williams, 2019). High level of autism traits = Social Responsiveness Scale-2 total *t*-score > 76 (Constantino, 2012).  ** = p < .05. ** p < .01.* | | | | | |

| **Table 7**  *Spearman's Correlations Coefficients of Services, Continuous Individual Differences, and Social-Ecological Factors* | | | | | | | | |
| --- | --- | --- | --- | --- | --- | --- | --- | --- |
|  | 1 | 2 | 3 | 4 | 5 | 6 | 7 | 8 |
| 1. Number of services | - |  |  |  |  |  |  |  |
| 2. Unmet service needs | -.00 | - |  |  |  |  |  |  |
| 3. Barriers to services | .17 | .50** | - |  |  |  |  |  |
| 4. CELF-5 core language score | -.40** | -.17 | -.18 | - |  |  |  |  |
| 5. NVIQ | -.42** | -.14 | -.08 | .59** | - |  |  |  |
| 6. SRS-2 total *t*-scores | -.07 | .45** | .27* | .09 | .00 | - |  |  |
| 7. Sense of community | .16 | -.43** | -.27* | -.00 | .12 | -.45** | - |  |
| 8. Educational enrollment: Yes | .39** | -.02 | -.00 | -.29* | -.23 | -.13 | .11 | - |
| *Note.* Barriers to services = barriers to having services needs met. CELF-5 = Clinical Evaluation of Language Fundamentals-5 Expressive Language Index (Wiig et al., 2013). NVIQ = nonverbal intelligence. SRS-2 = Social Responsiveness Scale-2nd Ed. (Constantino, 2012).  * = *p* < .05. ** = *p* < .01. | | | | | | | | |

| **Table 8**  *Negative Binomial Regression Results of Numbers of Services, Unmet Needs, and Barriers with Continuous Individual Difference Predictors* | | | | | | | | |
| --- | --- | --- | --- | --- | --- | --- | --- | --- |
|  | RR | 95% CI | *p* | | *χ^2^* | | *df* | *p* |
| *Number of Services* | | | | | | | | |
| Model |  |  | 20.18 | 3 | | < .001 | | |
| Intercept | 2.11** | 1.54, 2.88 |  |  | |  | | |
| NVIQ centered on 100 | 0.99 | .98, 1.00 |  |  | |  | | |
| CELF-5 core language score centered on 100 | .99 | .98, 1.00 |  |  | |  | | |
| Educational enrollment: No | 1.48* | 1.02, 2.15 |  |  | |  | | |
| *Unmet Service Needs* | | | | | | | | |
| Model |  |  | 15.21 | 2 | | < .001 | | |
| Intercept | 4.50** | 1.51, 13.44 |  |  | |  | | |
| SRS-2 total *t*-scores centered on 59 | 1.03* | 1.00, 1.05 |  |  | |  | | |
| Sense of community | 0.77 | .57, 1.05 |  |  | |  | | |
| *Barriers to Services* | | | | | | | | |
| Model |  |  | 5.38 | 2 | | .068 | | |
| Intercept | 6.32 | 3.23, 12.35 |  |  | |  | | |
| SRS-2 total *t*-scores centered on 59 | 1.01 | .10, 1.03 |  |  | |  | | |
| Sense of community | .91 | .77, 1.07 |  |  | |  | | |
| *Note.* RR = rate ratio. Clinical Evaluation of Language Fundamentals-5 (CELF-5) (Wiig et al., 2013), SRS-2 = Social Responsiveness Scale-2 total t-score > 76 (Constantino, 2012).  ** = p < .05. ** p < .01.* | | | | | | | | |
